# CSF proteome-wide study of neuropsychiatric symptoms of dementia

**DOI:** 10.64898/2026.06.26.26356698

**Authors:** Zhen Mei, Nicholas Howard, Danielle J. Harvey, Alzheimer’s Disease Neuroimaging Initiative, Edward Fox, Nicholas T. Seyfried, Thomas S. Wingo, Aliza P. Wingo

## Abstract

**Introduction:** Neuropsychiatric symptoms in dementia (NPS) are common and among the most troubling aspects of living with dementia, yet their underlying mechanisms remain unclear. Here, we aimed to identify cerebrospinal fluid (CSF) proteins associated with NPS.

**Methods:** Proteomes were profiled from CSF collected at baseline from participants of the Alzheimer’s Disease Neuroimaging Initiative (ADNI) using mass spectrometry. Here, we included participants having positive AD CSF biomarkers (i.e., pTtau181 / Aβ42 ratio >0.025) and mild cognitive impairment or AD dementia (n=419). Eight NPS domains were assessed longitudinally with the Neuropsychiatric Symptom Inventory Questionnaire. Severity of cognitive impairment was evaluated using the CDR-SB. Proteome-wide differential expression analysis for each NPS domain at baseline was performed. Significant protein-NPS associations underwent mediation analysis to test whether they were mediated by cognitive impairment severity. Cox proportional hazard was modeled for baseline CSF proteins and incident NPS. Additionally, we tested whether candidate NPS causal proteins previously identified in brain are associated with NPS in CSF.

**Results:** We identified 8 CSF proteins associated with apathy at baseline (FDR q<0.05) after adjusting for sex, age, and education - NTNG2, S100A1, FZD1, FSTL5, CDH7, CHODL, FBXO2, and CACNA2D2. Mediation analysis revealed these associations were independent of cognitive impairment severity in four proteins and only partially mediated by cognitive impairment severity in the remaining four proteins. Among the 10 NPS candidate causal proteins previously identified in brain and detected in CSF, the abundance of two proteins (CPD, GRN) in CSF was associated with baseline disinhibition and of two other proteins (PIK3IP1, PCMT1) with both baseline apathy and incident apathy after adjusting for sex, age, and education.

**Discussion:** These findings suggest that proteomic alterations in apathy in MCI/AD encompass synaptic connectivity, calcium regulation, Wnt-signaling, and neuronal proteostasis, highlighting potential CSF biological processes and biomarker candidates for apathy.

## Introduction

Neuropsychiatric symptoms in dementia (NPS) include a range of behavioral, psychological, and personality changes that are commonly experienced by individuals living with mild cognitive impairment (MCI) or Alzheimer’s disease (AD) or related dementia [1]. While NPS are not part of the diagnostic criteria for AD, epidemiological studies suggest that most individuals with MCI or AD experience one or more NPS, including apathy, depression, agitation, anxiety, irritability, sleep disturbances, disinhibition, and psychosis[2–5]. These symptoms are associated with poorer quality of life, greater functional and cognitive decline, and increased caregiver burden and earlier institutionalization[6, 7].

NPS are also linked to biomarkers of AD pathologies. The presence of NPS has been associated with lower cerebrospinal fluid (CSF) Aβ42 and higher p-tau and t-tau levels[8, 9]. In addition, the late-life emergence of NPS has been shown to correlate with increased *β*-amyloid deposition and higher risk of incident dementia among cognitively unimpaired individuals[10, 11], suggesting they may serve as both diagnostic and prognostic indicators of AD.

Studying protein correlates of NPS can inform disease mechanisms and guide early diagnosis and intervention. CSF is an informative biofluid for this purpose because it is in direct contact with the brain and can reflect neurological and neurodegenerative processes. Few studies have examined CSF proteins with altered expression levels in NPS. Using tandem mass tag (TMT)-based proteomics, one study of 87 participants reported 27 CSF proteins associated with composite NPS reflected by the total NPI-Q score [12]. Another study identified 57 CSF proteins associated with depressive symptoms in 688 individuals with cognitive decline or Alzheimer’s disease [13]. While both studies provide valuable information, a more comprehensive examination of each of the eight NPS domains is still lacking.

To fill this gap, we examined each of the eight NPS domains in 418 participants with MCI or AD dementia recruited by the Alzheimer’s Disease Neuroimaging Initiative study (ADNI). Using tandem mass tag (TMT) mass spectrometry, we profiled 2,961 proteins in CSF and examined them in each of the eight NPS domains. Additionally, following a hypothesis-driven approach, we examined the association between NPS and 10 NPS candidate causal proteins previously identified in brain proteogenomic studies and detected in CSF.

## Methods

### Study participants

We used phenotypic and CSF proteomic data of participants recruited by the Alzheimer’s Disease Neuroimaging Initiative study (ADNI) (adni.loni.usc.edu). Started in 2003, ADNI is a multicenter longitudinal study designed to track the progression of Alzheimer’s disease across normal aging, mild cognitive impairment (MCI), and AD dementia.

In ADNI, diagnostic criteria for AD include a Mini-Mental State Examination (MMSE) score between 20 and 26 and a global Clinical Dementia Rating (CDR) score of 0.5 or 1[14]. Criteria for MCI include a MMSE score between 24 and 30, a global CDR score of 0.5, a CDR memory score of at least 0.5, and objective memory impairment on Wechsler Memory Scale-Logical Memory II test[14]. The CDR Sum of Boxes (CDR-SB) was the sum of scores across all six CDR domains and was used as a continuous measure of cognitive impairment severity[15].

Since we investigate neuropsychiatric symptoms in MCI or dementia, our study included participants with MCI or AD dementia who had baseline CSF proteomic data. To enrich for participants with AD etiologies, we restricted our analyses to those (n=418) with CSF biomarkers for AD as reflected by pTau181/Aβ42 ratios >0.025, a cutoff that has been shown to be highly concordant with amyloid-β PET imaging in a prior study[16]. Here, CSF Aβ42 and pTau181 levels were measured using Roche Elecsys immunoassays. Given the low statistical power in other ethnic groups, our analysis was restricted to self-identified white participants.

### Neuropsychiatric symptoms

All domains of the neuropsychiatric symptoms in MCI or dementia (referred to as NPS here) were assessed longitudinally using the Neuropsychiatric Inventory-Questionnaire (NPI-Q). The NPI-Q assesses the presence and severity of neuropsychiatric symptoms across 12 domains, including delusions, hallucinations, agitation, depression, anxiety, euphoria/elation, apathy/indifference, disinhibition, irritability, aberrant motor behavior, sleep disturbances, and appetite/eating disturbances[17]. In ADNI-1/GO/2, NPI-Q was administered at baseline, 6, and 12 months, and annually thereafter; MCI participants in ADNI-1 also had an 18-month assessment. In ADNI-3, assessments were conducted annually.

For the present study, we focused on the 8 NPS that are commonly observed across the AD continuum including apathy, depression, anxiety, agitation, disinhibition, irritability, sleep disturbances and psychosis (defined as the presence of delusions and/or hallucinations)[2, 3]. Participants were classified based on the presence or absence of symptom for each NPI domain. All participants included in our analysis had baseline NPS assessment and at least two follow-up NPS assessments.

### Cerebrospinal fluid proteomic data

CSF was collected at baseline and proteins were extracted for proteomic sequencing using tandem mass tag mass spectrometry. Methods for CSF proteomic sequencing have been described in detail previously[18]. In brief, baseline CSF samples from ADNI participants were first digested and labeled with TMTpro reagents. A global internal standard (GIS) was created to normalize protein quantification across batches. Samples were fractionated by high-pH reversed-phase chromatography and then analyzed by liquid chromatography coupled to tandem mass spectrometry. The raw files (Synapse SynID: syn59804727) were searched using FragPipe v22.0 against a UniProt human protein database with default TMT-16 plex parameters. Peptide Spectral Matches were validated using Percolator, with the false discovery rate (FDR) threshold of 1%. Protein and peptide abundances were quantified using Philosopher. The data from all batches were merged and protein levels were first scaled by dividing each protein intensity by sum of intensity of all proteins in each sample, followed by multiplying by the maximum protein intensity sum across all samples. Instances where the intensity was ‘0’ were treated as ‘missing values’.

For quality control of proteomic data, we kept proteins with protein abundance measurable in ≥ 200 subjects and selected the highest abundance entry for proteins with multiple UniProt annotations. Each protein’s abundance was normalized by dividing it by the sum abundance of all the proteins profiled for that sample, followed by the log2 transformation. To assess potential blood contamination, hemoglobin subunit levels were examined but no outlier samples were detected, suggesting no blood contamination. We excluded samples with > 4 standard deviations from the mean of the first two principal components using iterative PCA. Linear regression was used to regress out the effect of protein sequencing batch, and the residual protein abundance was used for downstream analyses. After quality control, there were 2961 proteins in the CSF proteomes for downstream statistical analysis.

### Identification of candidate NPS causal proteins in brain

With robust evidence from large-scale human data – epidemiologic studies[1, 19–24], GWAS, brain transcriptomic, and brain proteomic studies[25–29] – we posit that shared causal proteins between AD and psychiatric disorders underlie NPS in MCI/AD. We previously identified putative causal proteins for each of the 16 major psychiatric disorders and 6 neurodegenerative diseases through integration of GWAS with brain proteogenomic data for each condition using proteome-wide association study (PWAS) and Mendelian randomization[28]. We identified 1140 candidate causal proteins for these 21 conditions. We observed that a third of the identified candidate causal proteins in AD were also causal proteins in one or more psychiatric disorders. These shared causal proteins between AD and psychiatric disorders are referred to as candidate NPS brain causal proteins (*n*=22, Table S1)[28]. Of these 22 proteins, 10 were detected in CSF proteomes, including ACE, CPD, CPLX1, GC, GRN, LPP, PCMT1, PIK3IP1, STX1B, and TMEM106B (Table S1). In the present study, we examined these 10 proteins in relation to baseline and incident NPS domains.

### Statistical analysis

All statistical analyses were performed in R (v4.4.0). We first compared baseline clinical characteristics between participants with MCI and AD dementia. Continuous variables (e.g., CDR-SB) were compared using linear regression models, and binary variables (e.g., NPI domains) were compared using logistic regression models. All models were adjusted for sex, age, and years of education.

Proteome-wide differential expression analysis for each of the eight NPS domains was performed with protein abundance as the outcome and baseline NPS as the independent variable. All models were adjusted for sex, age, and education, with Benjamini-Hochberg false discovery rate (FDR) correction applied across all proteins.

Mediation analyses for those NPS associated with protein abundance were conducted in R using the mediation package[30], with protein as the treatment variable, NPS as the outcome, and cognitive impairment severity assessed by CDR-SB as the mediator. Both the mediator and outcome models were adjusted for sex, age, and education. As protein is a continuous variable, we set the treated and control conditions as the mean + 1 SD and the mean expression level, respectively, for estimation of the mediation effects. p-values and 95% confidence intervals were estimated based on 10,000 bootstrapped resamples. Mediation was considered significant when the average causal mediation effect (ACME) was significant (p < 0.05), and independent of the mediator otherwise. For significant mediation effects, mediation was classified as complete if the direct effect was non-significant, or partial if the direct effect remained significant (p < 0.05) and 95% confidence interval for the proportion mediated did not include 1.

Cox proportional hazard modeling was performed to determine if proteins that are associated with NPS at baseline are also associated with the hazard of developing NPS during follow-up. In the primary analyses, only participants without NPS at baseline were included and Cox models were adjusted for sex, age, and education. The event was defined as the first occurrence of NPS during follow-up. Participants who did not develop NPS were censored at their last follow-up visit. As a sensitivity analysis, we included all participants in the Cox models and additionally adjusted for baseline NPS, as well as sex, age, and education. Schoenfeld residuals tests were used to evaluate the proportional hazards assumption in Cox models.

## Results

### Participant characteristics

A total of 418 participants were included in our analysis (**Table 1**), all of whom had baseline and longitudinal NPS assessments, baseline CSF proteomic data, and positive CSF pTau181/Aβ42 ratios (ratio >0.025). Of these participants, the mean age at baseline was 73.9 years, 38% were female, and the mean years of education was 15.8.

**Table 1.** Participant characteristics.

|  | **All** | **MCI<sup>a</sup>** | **AD<sup>a</sup>** |
| --- | --- | --- | --- |
| No. of participants (%) | 418 | 280 (67%) | 138 (33%) |
| Sex, female (%) | 159 (38%) | 100 (35.7%) | 59 (42.8%) |
| Age at baseline, y, mean (SD) | 73.9 (7.54) | 73.7 (7.24) | 74.2 (8.14) |
| Years of education, mean (SD) | 15.8 (2.83) | 15.9 (2.82) | 15.6 (2.85) |
| CDR-SB, mean (SD) | 2.53 (1.73) | 1.64 (0.917) | 4.32 (1.60) |
| APOE ε4, No. of carriers (%) | 296 (70.8%) | 194 (69.3%) | 102 (73.9%) |
| CSF Aβ1-42, pg/mL, mean (SD) | 640 (217) | 669 (224) | 579 (188) |
| CSF p-tau181, pg/mL, mean (SD) | 35.5 (13.5) | 35.0 (13.3) | 36.4 (14.0) |
| Participants with NPS at baseline, n (%) |  |  |  |
| Agitation | 89 (21.3%) | 51 (18.2%) | 38 (27.5%) |
| Anxiety | 87 (20.8%) | 44 (15.7%) | 43 (31.2%) |
| Apathy | 100 (23.9%) | 51 (18.2%) | 49 (35.5%) |
| Depression | 119 (28.5%) | 69 (24.6%) | 50 (36.2%) |
| Disinhibition | 49 (11.7%) | 26 (9.3%) | 23 (16.7%) |
| Irritability | 120 (28.7%) | 76 (27.1%) | 44 (31.9%) |
| Psychosis | 24 (5.7%) | 11 (3.9%) | 13 (9.4%) |
| Sleep disturbance | 75 (17.9%) | 44 (15.7%) | 31 (22.5%) |
| Follow-up years, mean (SD), max | 3.71 (2.74), 16.5 | 4.59 (2.86), 16.5 | 1.93 (1.15), 10.8 |
| Follow-up visits, median (min, max) | 4 (2, 16) | 6 (2, 16) | 3 (2, 7) |

a. All MCI and AD participants had abnormal CSF pTau181/Aβ42 ratios (i.e., ratio >0.025).

At baseline, 280 participants (67%) had MCI and 138 (33%) had AD dementia (**Table 1**). As expected, participants with AD dementia had higher CDR-SB (*β* = 2.7; *p*<0.001) and lower CSF Aβ1-42 levels (*β* = −96.3; *p*<0.001) compared to those with MCI, while there were no differences in APOE ε4 status (*p*=0.27) and CSF p-tau181 levels (*p*=0.41). The odds ratios for experiencing NPS were significantly higher in AD dementia compared to MCI in six NPS domains (Table S2), suggesting a positive association between cognitive impairment and NPS.

All participants included in our analysis had at least two longitudinal NPS assessments (**Table 1**). The mean follow-up time was 3.71 years, with a maximum follow-up of 16.5 years.

### Associations between baseline CSF proteins and baseline NPS

To identify CSF proteins associated with NPS manifesting at baseline, we performed a proteome-wide differential expression analysis for each of the 8 NPS domains, adjusting for sex, age, and education. We found 8 proteins associated with apathy in MCI / AD participants (FDR q<0.05, **Figure 1a**, Table S3), while no proteins reached FDR significance for other NPS domains (Table S4-S10). These 8 proteins included NTNG2, S100A1, FZD1, FSTL5, CDH7, CHODL, FBXO2, and CACNA2D2 (Figure 1a). Existent literature suggests they are primarily involved in synaptic connectivity[31–36], calcium regulation [37–40], Wnt-signaling[41], and neuronal proteostasis[42].

**Figure 1:**
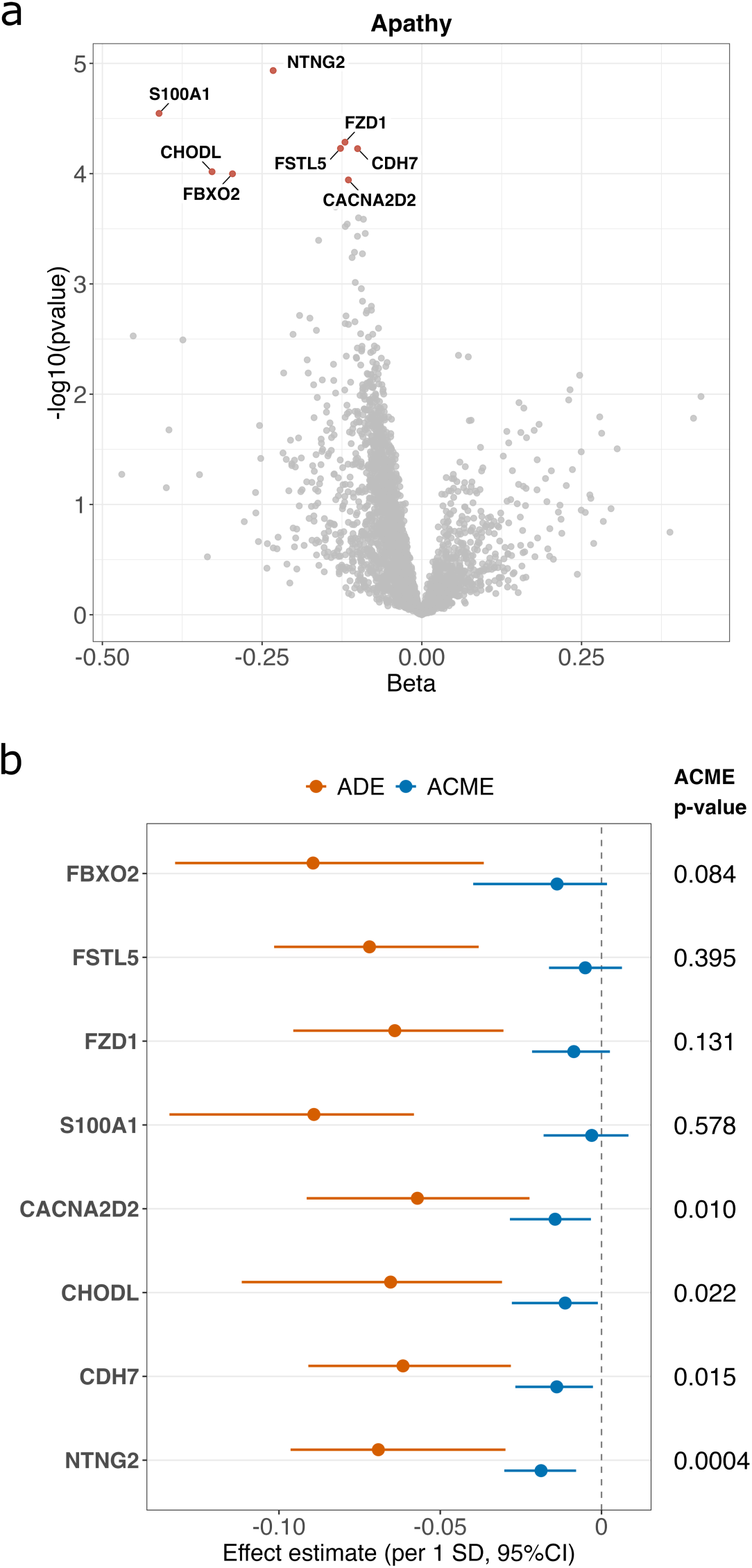
CSF proteins differentially expressed in NPS. (a) The volcano plot showing results of the proteome-wide differential analysis of apathy, adjusting for sex, age, and education. The x-axis depicts the beta coefficients and the y-axis depicts the −*log*_10_(*p values*) from linear regression modeling of each protein and apathy, adjusting for sex, age, and education. The red points indicate significant proteins at FDR q<0.05. Beta coefficients and statistics for all proteins are provided in the Table S3. (b) Results of the mediation analysis where the severity of cognitive impairment was the mediator. Average causal mediation effects (ACME) and average direct effects (ADE) were estimated using the R package mediation with protein as the treatment variable, apathy as the outcome, and CDRSB as the mediator. Significant mediation is reflected by ACME p-value<0.05 or a 95% confidence interval (CI) that does not include 0. In the case of significant mediation, partial mediation is defined by an ADE p-value<0.05 or a 95% CI that does not include 0.

To examine if cognitive impairment explains the associations between these proteins and apathy, we performed mediation analysis with protein as the treatment variable, apathy as the outcome, and CDR-SB as the mediator. Four proteins (NTNG2, CACNA2D2, CDH7, and CHODL) showed partial mediation through CDR-SB (**Figure 1b**, Table S11), with an average mediation proportion of about 21% for NTNG2, 20% for CACNA2D2, 18% for CDH7, and 15% for CHODL (Table S11). No significant mediation by cognitive impairment severity was observed for the remaining four proteins (S100A1, FBXO2, FSTL5, and FZD1), suggesting their direct associations with apathy independent of the degree of severity of cognitive impairment (**Figure 1b**, Table S11).

### Associations between candidate NPS causal proteins and baseline NPS

Following a hypothesis driven approach, we examined associations between each of eight NPS domains and the 10 NPS candidate causal proteins previously identified in GWAS-informed brain proteogenomic studies and detected in CSF (see methods, Table S1).

In linear regression models adjusting for sex, age, and education, we found two NPS candidate causal proteins (PIK3IP1, PCMT1) differentially expressed in apathy at baseline (p <0.05; **Figure 2a**, Table S12) and another two proteins (CPD, GRN) differentially expressed in disinhibition at baseline (p<0.05; **Figure 2b**, Table S12). These findings suggest candidate NPS causal proteins from brain proteogenomic studies also showed differential expression in CSF.

**Figure 2:**
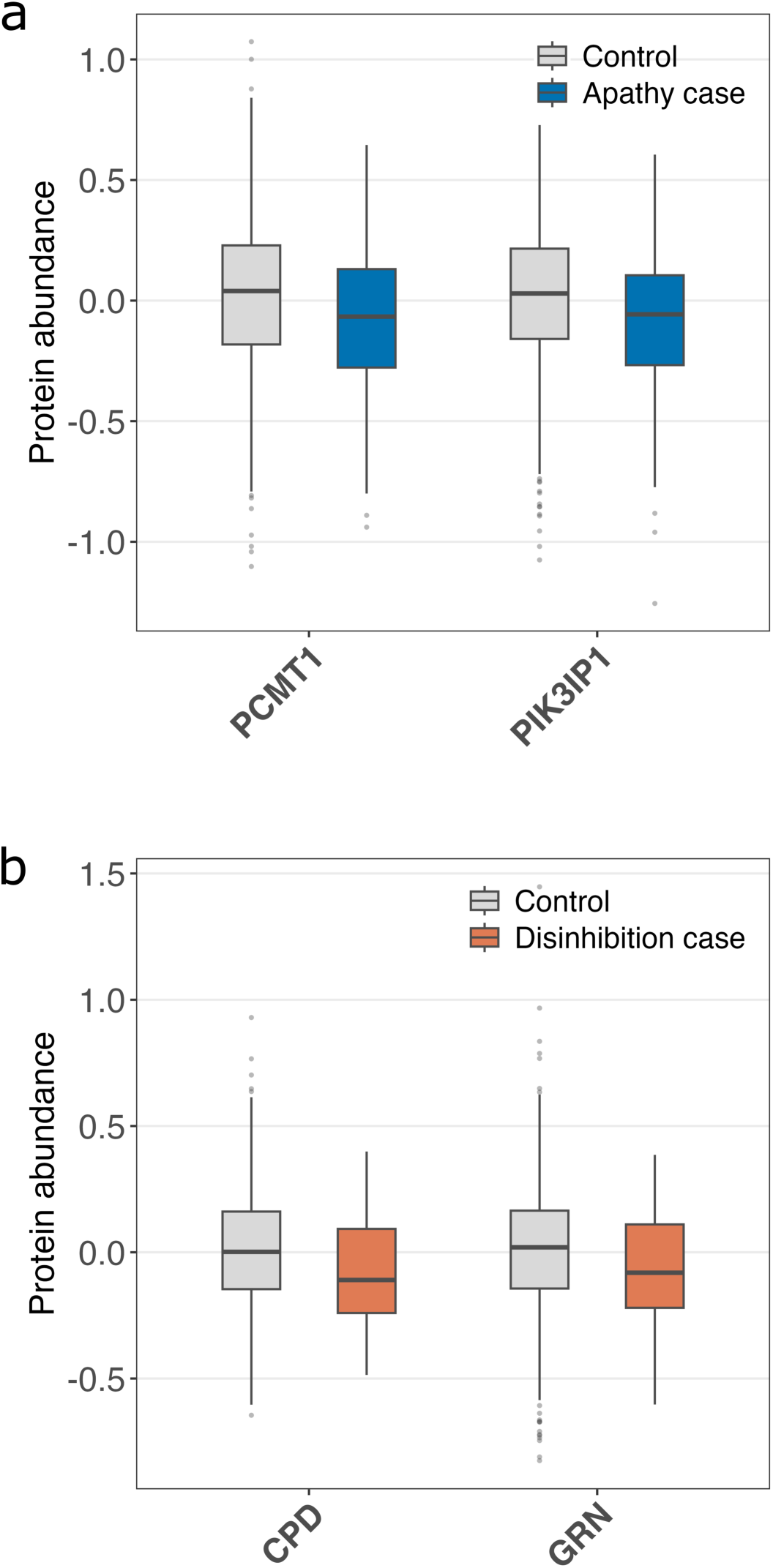
Associations between baseline NPS and candidate NPS causal proteins identified in brain and detected in CSF. Candidate NPS causal proteins differentially expressed in **(a)** apathy and **(b)** disinhibition. Boxplots display the median, 25th, and 75th percentiles, with whiskers (error bars) extending to the last data point within 1.5 times the interquartile range from the lower and upper quantiles.

### Associations between CSF proteins and the incident NPS

To examine whether the 12 proteins that were associated with NPS at baseline are also associated with the risk of developing NPS in follow-up visits, we performed Cox proportional hazards modeling among participants without NPS at baseline. The event of interest was defined as the first occurrence of NPS during follow-up. In the primary model adjusting for sex, age, and education, we found that lower expression of four proteins was each associated with higher risk of apathy at *p*<0.05 for PIK3IP1 (HR = 0.43), CACNA2D2 (HR = 0.47), PCMT1 (HR = 0.56), and CHODL (HR = 0.59) (**Figure3,** Table S13). Cumulative hazard plots for these proteins and incident apathy were presented in **Figure 3**. CPD (*p*=0.07) and GRN (*p*=0.06) were not significantly associated with incident disinhibition (Table S14).

**Figure 3:**
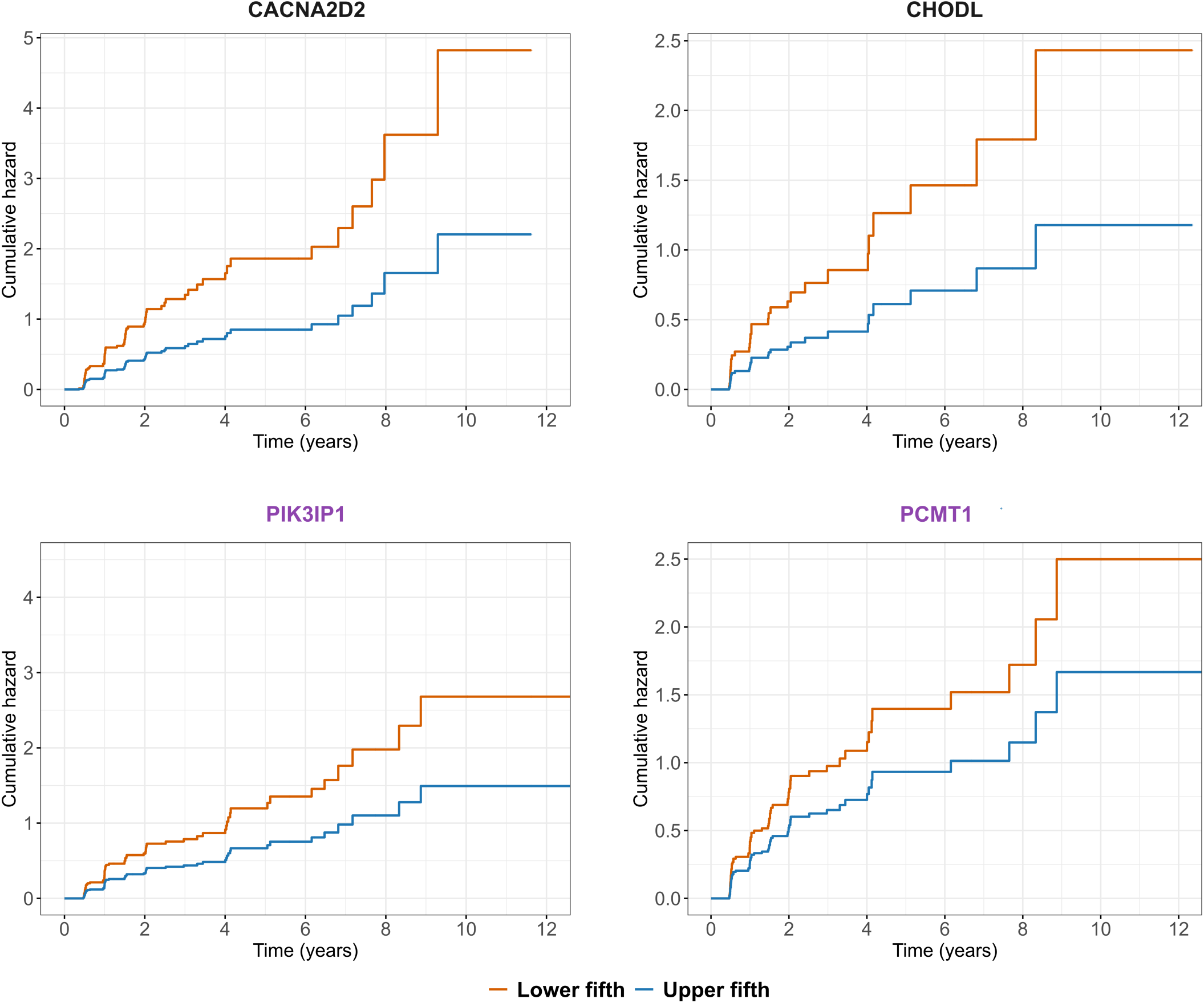
Associations between baseline CSF protein abundance and incidence of apathy. Cox proportional hazard regression was performed for baseline CSF proteins and incident NPS adjusting for sex, age, and education. Cumulative hazard plots illustrate associations between a protein measured at baseline and future incidence of apathy. Depicted are protein levels in participants in the lowest versus highest quintile for protein expression. Protein names in black are CSF proteins that were differentially expressed in apathy at baseline. Protein names in purple are candidate NPS causal proteins that were associated with apathy at baseline. Full results are provided in Table S13.

As a sensitivity analysis, we included all the participants (n=418) in the Cox proportional hazards model and additionally adjusted for baseline apathy with sex, age, and education. Three of four proteins remained significantly associated with the risk of incident apathy (Table S13), suggesting the robustness of our findings.

## Discussion

This is the first study that examines CSF proteins in each of the eight NPS domains in biomarker-confirmed MCI/AD participants to the best of our knowledge. We found eight proteins associated with apathy at a stringent significance threshold (FDR<0.05) and two of them (CACNA2D2, CHODL) were also associated with incident apathy. Mediation analysis showed associations for four proteins were independent of cognitive-impairment severity and the associations of the remaining four proteins were partially mediated by it. These eight apathy-associated proteins encompass synaptic connectivity (NTNG2, CDH7, CHODL)[31–36], calcium-related processes (CACNA2D2, S100A1, FSTL5)[37–40], Wnt-mediated synaptic maintenance (FZD1)[41], and neuronal proteostasis (FBXO2)[42].

Furthermore, following a hypothesis driven approach, we found two brain candidate NPS causal proteins (GRN, CPD) differentially expressed in CSF of participants with baseline disinhibition, and two other proteins (PIK3IP1, PCMT1) that were associated with apathy at baseline as well as with incident apathy. GRN acts as a critical regulator of lysosomal function and GRN variants are associated with both AD[43] and bipolar disorder[44]. PIK3IP1 is a negative regulator of the PI3K pathway and has been linked to both AD [45] and schizophrenia [46]. PCMT1 is a protein repair enzyme that plays a neuroprotective role in AD by inhibiting isoaspartate accumulation in AD-related proteins[47, 48]. Robust evidence from large-scale human studies suggests shared mechanisms and overlapping biological pathways between AD and psychiatric disorders[25–29]. Targeting these shared mechanisms may treat psychiatric disorders and simultaneously reduce dementia risk (e.g., lithium treatment)[49–51]. Our results further suggest that these shared risk proteins could be differentially expressed in NPS and may serve as CSF biomarkers for NPS in the context of MCI and AD.

Some apathy-associated proteins were involved in calcium regulation. CACNA2D2 encodes the α2δ subunit of voltage-gated calcium channels (VGCCs), which has a major role in trafficking the VGCCs to specific neuronal membrane domains[37, 38]. Genetic studies have linked CACNA2D2 variants to intellectual disability and schizophrenia[52, 53]. Our previous work also identified CACNA2D2 as a candidate causal protein for major depression, neuroticism, and tobacco use[28]. Besides CACNA2D2, S100A1 functions as a calcium sensor protein[39]; CDH7 mediated calcium-dependent cell adhesion[34]; and FSTL5 is a secreted protein with a predicted calcium-binding domain[40]. These findings suggest dysregulated calcium signaling may underlie apathy in MCI and AD.

Synaptic connectivity was another functional theme identified among the apathy-associated proteins. NTNG2[31–33], CDH7[34], and CHODL[35, 36] are all cell-surface proteins that regulate formation and specificity of neuronal connections. Genetic studies have linked CDH7[54–56] and NTNG2[57] to psychiatric disorders such as schizophrenia, bipolar disorder, and major depression. Although CHODL has been less studied in the context of psychiatric traits, its association with both baseline and incident apathy in the present study suggests it may be a candidate CSF biomarker that warrants further investigation.

The findings that the apathy-associated proteins were independent or partially independent from severity of cognitive impairment in the mediation analysis support the notion that apathy has a distinct molecular biology from the general synaptic loss driving cognitive decline and impairment in AD. CSF proteomics is well-suited for studying NPS biology since CSF is in direct contact with the brain tissue and contains proteins released from the brain, which reflect neurodegeneration, active secretion from glia, and proteolytic processing. CSF proteomics reveals the molecular processes active at the time of NPS assessment.

PIK3IP1 and PCMT1 were previously identified as candidate brain causal protein in NPS and are both differentially expressed in CSF in apathy and predicts future apathy. Baseline CSF PIK3IP1 and PCMT1 levels could potentially enrich clinical trials of apathy-targeted therapies analogous to how amyloid PET or CSF Abeta42 enriches anti-amyloid trials.

Our study has several strengths. First, we performed CSF protein differential expression analysis in each of the eight NPS domains at a stringent significance threshold (FDR<0.05) using deep MS-based proteomics. Our findings provide biological insights into specific domains of NPS rather than the general burden of NPS. Second, we examined associations between candidate NPS causal proteins from brain proteogenomic studies and NPS in CSF. Third, we performed mediation analysis to quantify the extent to which cognitive-impairment severity explains the associations between the protein and NPS. Fourth, we restricted our analyses to biomarker-confirmed AD participants, anchoring our findings in confirmed pathology. Lastly, consistent with a transcriptomic study of post-mortem anterior cingulate cortex in NPS[58], we found downregulated expression patterns for most NPSs with nominal p<0.05. Moreover, three proteins (FBXO2, FSTL5, NTNG2) were also associated with apathy in the study of postmortem brain (p<0.05)[59], supporting the validity of our protein models.

Our results should be interpreted in the context of the study’s limitations. First, among 8 tested NPSs, we only found differentially expressed proteins for apathy with an FDR<0.05, which may imply a lack of statistical power or relatively small effect sizes. Future studies with larger sample sizes or combining results from different cohorts may increase the ability to detect altered proteins in NPS. Second, NPI-Q used in this study was completed by the informants or caregivers, which may be potentially influenced by the recall bias. Third, our analysis was based on self-identified white participants owing to low statistical power in other groups, which potentially limits the generalizability of our findings.

In conclusion, our study identified some CSF proteins associated with baseline NPS and the incident risk. These findings provide some novel insights for future biomarker study of NPS.

## Data Availability

The data used in this study were obtained from the Alzheimer’s Disease Neuroimaging Initiative (ADNI) database. The data are available to qualified investigators through the ADNI data access process and can be obtained after registration and approval via the ADNI website: adni.loni.usc.edu.

## Acknowledgements

The authors are grateful to the ADNI participants who made this research possible. This work was supported by Veterans Affairs I01 BX003853 (A.P.W.), Veterans Affairs I01 BX005686 (A.P.W.), Veterans Affairs IK4 BX005219 (A.P.W.), National Institutes of Health R01 AG056533 (A.P.W., T.S.W.), National Institutes of Health R01 AG072120 (A.P.W., T.S.W.), National Institutes of Health R01 AG075827 (A.P.W., T.S.W.), National Institutes of Health R01 AG079170 (T.S.W.).

Data collection and sharing for the Alzheimer’s Disease Neuroimaging Initiative (ADNI) is funded by the National Institute on Aging (National Institutes of Health Grant U19 AG024904). The grantee organization is the Northern California Institute for Research and Education. In the past, ADNI has also received funding from the National Institute of Biomedical Imaging and Bioengineering, the Canadian Institutes of Health Research, and private sector contributions through the Foundation for the National Institutes of Health (FNIH) including generous contributions from the following: AbbVie, Alzheimer’s Association; Alzheimer’s Drug Discovery Foundation; Araclon Biotech; BioClinica, Inc.; Biogen; Bristol-Myers Squibb Company; CereSpir, Inc.; Cogstate; Eisai Inc.; Elan Pharmaceuticals, Inc.; Eli Lilly and Company; EuroImmun; F. Hoffmann-La Roche Ltd and its affiliated company Genentech, Inc.; Fujirebio; GE Healthcare; IXICO Ltd.; Janssen Alzheimer Immunotherapy Research & Development, LLC.; Johnson & Johnson Pharmaceutical Research &Development LLC.; Lumosity; Lundbeck; Merck & Co., Inc.; Meso Scale Diagnostics, LLC.; NeuroRx Research; Neurotrack Technologies; Novartis Pharmaceuticals Corporation; Pfizer Inc.; Piramal Imaging; Servier; Takeda Pharmaceutical Company; and Transition Therapeutics.

